# Clinical phenotypes and prognostic features of ETMRs (Embryonal Tumor with Multi-layered Rosettes) a new CNS tumor entity: A Rare Brain Tumor Registry study

**DOI:** 10.1101/2020.08.12.20171801

**Authors:** Sara Khan, Palma Solano-Paez, Tannu Suwal, Mei Lu, Salma Al-Karmi, Ben Ho, CV AlmeidaGonzalez, Derek Stephens, Andrew Dodgshun, Mary Shago, Paula Marrano, Adriana Fonseca, Lindsey M. Hoffman, Sarah Leary, Holly B. Lindsay, Alvaro Lassaletta, Anne E. Bendel, Christopher Moertel, Andres Morales, Vicente Santa-Maria, Cinzia Lavarino, Eloy Rivas, Sebastian Perreault, Benjamin Ellezam, Nada Jabado, Angelica Oviedo, Michal Yalon-Oren, Laura Amariglio, Helen Toledano, James Loukides, Timothy E. Van Meter, Hideo Nakamura, Tai-Tong Wong, Kuo-Sheng Wu, Chien-Jui Cheng, Young-Shin Ra, Milena La Spina, Luca Massimi, Anna Maria Buccoliero, Alyssa Reddy, Rong Li, G. Yancey Gillespie, Dariusz Adamek, Jason Fangusaro, David Scharnhorst, Joseph Torkildson, Donna Johnston, Jean Michaud, Lucie Lafay-Cousin, Jennifer Chan, Frank Van Landeghem, Beverly Wilson, Sandra Camelo-Piragua, Nabil Kabbara, Mahjouba Boutarbouch, Derek Hanson, Chad Jacobsen, Karen Wright, Jean M. Mulcahy Levy, Yin Wang, Daniel Catchpoole, Nicholas Gerber, Michael A. Grotzer, Violet Shen, Ashley Plant, Christopher Dunham, Maria Joao Gil da Costa, Ramya Ramanujachar, Eric Raabe, Jeffery Rubens, Joanna Philips, Nalin Gupta, Ahmet Demir, Christine Dahl, Mette Jorgensen, Eugene I. Hwang, Amy Smith, Enrica Tan, Sharon Low, Jian-Qiang Lu, NG Ho-Keung, Jesse L Kresak, Sridharan Gururangan, Scott L. Pomeroy, Nongnuch Sirachainan, Suradej Hongeng, Vanan Magimairajan, Roona Sinha, Naureen Mushtaq, Reuben Antony, Mariko Sato, David Samuel, Michal Zapotocky, Samina Afzal, Nisreen Amayiri, Maysa Al-Hussaini, Andrew Walter, Tarik Tihan, Gino R. Somers, Amar Gajjar, Paul Wood, Nicolas Gottardo, Jason E. Cain, Peter A Downie, Helen Branson, Suzanne Laughlin, Brigit Ertl-Wagner, Derek S. Tsang, Vijay Ramaswamy, James Drake, Abhaya V. Kulkarni, David S Ziegler, Sumihito Nobusawa, Uri Tabori, Michael D. Taylor, George M Ibrahim, James T. Rutka, Peter B. Dirks, Lili-Naz Hazrati, Richard G. Grundy, Maryam Fouladi, Pr Laetitia Padovani, Franck Bourdeaut, Jordan R. Hansford, Ute Bartels, Christelle Dufour, Cynthia Hawkins, Nicolas Andre, Eric Bouffet, Annie Huang

## Abstract

**Background:** ETMRs are a newly recognized rare paediatric brain tumor with alterations of the *C19MC* microRNA locus. Due to varied diagnostic practices and limited clinical data, disease features and determinants of outcome are poorly defined. We performed an integrated clinico-pathologic and molecular analyses of 159 primary ETMRs to define clinical phenotypes, identify predictors of survival and critical treatment modalities for this orphan disease.

**Methods:** Primary ETMR patients were identified from the Rare Brain Tumor Consortium (rarebraintumorconsortium.ca) global registry using histopathologic and molecular assays. Event-Free (EFS) and Overall Survival (OS) for 108 patients treated with curative multi-modal regimens were determined using Cox proportional hazard and log rank analyses.

**Findings:** ETMRs were predominantly non-metastatic (73%) tumors arising from multiple sites; 55% were cerebral tumors, 45% arose at sites characteristic of other brain tumors. Hallmark *C19MC* alterations were seen in 91%; 9% were ETMR-NOS. Survival and hazard analyses showed a 6 month median EFS and 2-4yr OS of 27-29% with metastatic disease (HR=0.44, 95% CI 0.26-0.74; p=0.002) and brainstem location (HR=0.40, 95% CI 0.021-0.75; p=0.005) correlating with adverse OS. Gross total resection (GTR: HR=0.38, 95% CI 0.21-0.68; p=0.001), high dose chemotherapy (HDC: HR=0.55, 95% CI 0.31-0.97; p=0.04) and radiation (RT: HR=0.32, 95% CI 0.16-0.60; p=<0.001) correlated with improved EFS and OS in multi-variable analyses. EFS and OS for patients treated with only conventional dose chemotherapy (CC) was 0% and respectively 37%±14% and 32%± 13% for patients treated with HDC. Patients with GTR or sub-total resection (STR) treated with HDC and RT had superior EFS (GTR 73%±14%, p=0.018; STR 67%±19% p=0.009) and OS (GTR 66%±17%, p=0.05; STR 67%±16%, p=0.005). Amongst 21 long-term survivors (OS 24-202 months); 38%, 24% and 24% respectively received craniospinal, focal or no RT.

**Interpretation:** Prompt molecular diagnosis and post-surgical treatment with multi-modal therapy tailored to patient-specific risk features improves ETMR survival.

**Funding:** This work was supported by the Canadian Institute of Health Research Grant No. 137011, Canada Research Chair Awards to AH. Funds from Miracle Marnie, Phoebe Rose Rocks, Tali’s Funds, Garron Cancer Centre, Grace’s Walk, Meagan’s Walk, Nelina’s Hope and Jean Martel Foundation are gratefully acknowledged. SK and PS were respectively supported by the Australian Lions Children’s Cancer Foundation and the Spanish Society of Pediatrics, Consejería de Salud y Familias de la Junta de Andalucía Project EF-0451-2017.

## INTRODUCTION

Embryonal tumors represent the largest category of brain cancers arising in children 0-14 years of age (1). Although medulloblastoma is most common, 40% of pediatric embryonal brain tumors are rare entities primarily affecting very young children and including Rhabdoid tumors and tumors historically categorized as supra-tentorial or CNS primitive neuroectodermal brain tumors (2, 3). Recent global profiling studies show these rare tumors span a spectrum of molecular diseases leading to the designation of new WHO diagnostic categories (2, 4-7). However, as many of these entities were historically considered and treated as different diseases, existing clinical data is difficult to evaluate. Furthermore, as molecular diagnoses of these rare diseases is not routinely performed, the clinical and therapeutic spectrum of these new, rare diseases remain poorly defined. Delays in clinical recognition, diagnosis and commencement of specific treatment likely contributes significantly to the generally poorer outcome of children with rare brain tumors.

Eberhart et al, first described a new histologic category of infant brain tumors, called Embryonal Tumor with abundant Neuropil and True Rosettes (ETANTR) (8). Subsequent studies showed various rare histologic entities, considered to be distinct diagnoses including ETANTR, Medullo-epithelioma (MEP), Ependymoblastoma (EPB) and supra-tentorial Primitive Neuroectodermal Tumors (sPNET), comprised a common molecular entity defined by recurrent amplification/gene fusions of *C19MC*, a novel oncogenic microRNA locus and primitive transcriptional and epigenetic features enriched for LIN28A, a pluripotency factor (4-7, 9). These studies led to the designation *C19MC*-altered ETMRs and a small proportion of tumors without *C19MC* alterations (ETMR NOS), as a new WHO CNS diagnostic category (2). While FISH analyses specifically identifies *C19MC*-altered ETMRs, ETMR NOS are more challenging diagnoses due to LIN28A expression in other malignant pediatric brain tumors including Rhabdoid tumors, high grade gliomas, germ cell tumors (6, 10-12), as well as non-CNS cancers (13).

ETMRs are now increasingly reported in the literature indicating this previously under-recognized entity may comprise a significant proportion of embryonal brain tumors arising in infants and younger children. Due to the varied histologic labels and relatively recent discovery of a molecular diagnostic marker in 2009, there is limited to no prospective treatment and outcome data on ETMRs. Due to early studies indicating ETMRs are lethal diseases with <10% overall survival (4, 6) ETMR patients are frequently treated with various aggressive conventional multi-modal and experimental therapies. Although benefits of high dose chemotherapy and radiation has been reported in various small studies and literature reviews, the impact of these treatments (14-16), have been difficult to assess due to limited patient cohorts, inconsistent diagnostic methods for ETMRs and lack of treatment and outcome information for the majority of patients reported to date. In order to inform current clinical practice and future clinical trials, we examined molecular, clinical, treatment and survival features of a large cohort of primary ETMRs patients identified from the Rare Brain Tumor clinical registry (rarebraintumorconsortium.ca) using multiple diagnostic assays.

## MATERIALS AND METHODS

### Patient, sample selection and procedures

The Rare Brain Tumor Consortium comprised of 140 participating centres, was established in 2002 to generate a comprehensive clinical database and biorepository for rare pediatric brain tumors. Tumor samples and clinical information were collected from treating physicians with patient consent as per Research Ethics Board guidelines at the Hospital for Sick Children and all participating institutions (Table S1). For this study clinical and diagnostic materials from 1904 patients in the RBTC database were reviewed to identify patients with institutional diagnoses of embryonal brain tumors; 99 patients with insufficient tissue were excluded. Based on review of histopathologic data, 213 tumors (Table S2) were further examined by multiple molecular assays as described in section below (Table S3). Data from 159 patients with a confirmed molecular diagnosis of ETMR were reviewed for age, sex, tumor location, metastatic status as defined by the Chang staging system, treatment received (extent of surgery, type of chemotherapy regimen, radiation field and dose), event free and overall survival. Only patients with complete clinical and treatment data were included in clinicopathologic and prognostic analyses. Patients who received a combination of oncologic tumor surgery, multi-agent chemotherapy with or without radiation, regarded globally as standard approach for embryonal brain tumors, were considered as treated with curative intent. Patients treated with surgery alone or lacking details of chemotherapy were not considered as treated with curative intent. Detailed molecular, clinical and outcome data current to November 2019 (Table S2, S3) was used for all correlative analyses.

### Molecular and immuno-histochemical assays

DNA/RNA from frozen or formalin-fixed, paraffin-embedded tumor tissue were processed for RNA sequencing or profiling on the Illumina Human 450K or EPIC methylation arrays (Illumina, San Diego, CA) as previously described (6). *C19MC* alterations were confirmed using a combination of SNP and methylation array derived copy number analyses (Conumee program; version 1.8.0) (Table S3) and C19MC FISH using test (RP11-381E3) and control (RP11-451E20) probes as previously described (4), while gene fusions were identified using Illumina RNA sequencing. To identify ETMRs NOS, global methylation data from 1805 tumors were examined relative to an in house reference data set of 1200 pediatric brain tumors with established diagnoses, using multiple orthogonal analyses. A t-distributed stochastic neighbor embedding (tSNE; Rtsne v0.15) analyses, based on the 12,500 most variable methylation probes determined by standard deviation (SD), was used to visualize global methylation features of ETMRs relative to the reference pediatric brain tumor data set. Reagents, methods used for LIN28A IHC staining and scoring (ML, TS) were as in prior publications (6).

### Statistical and informatic analyses

Overall (OS) and Event-free survival (EFS) were respectively defined as time from diagnosis to disease-related death or last visit, and time from diagnosis to first progression, relapse, death or date of last visit. Univariate and multivariate Cox proportional hazard analyses were used to identify clinical, molecular or treatment prognostic factors. Age, tumor location and *C19MC* status were respectively considered continuous and categorical variables, while sex, metastatic status, gross total (GTR) or sub-total (STR) tumor resection, high (HDC) or conventional dose chemotherapy (CC) and radiation treatment were considered dichotomous variables. Effect of chemotherapy, radiation and surgical resection were analyzed using Cox-regression for time-dependent covariates. All tests were two-sided; p <0.05 was considered statistically significant. Kaplan-Meier method and log-rank tests were performed for comparative survival analyses using SPSS and R (v25. NY:IBM Corp; v3.6.1).

## RESULTS

### Histopathologic and molecular characteristics

We identified 159 primary and 15 recurrent samples from patients enrolled on the RBTC registry that met histologic and molecular diagnostic criteria for ETMRs; only data from primary ETMRs were included in clinico-pathologic analyses.

Consistent with the unique identity of ETMRs, cluster analyses of global methylation profiles showed C19MC-altered ETMRs and ETMR NOS segregated from other pediatric CNS tumors (Figure 1A). Although *C19MC* amplification has been most commonly reported in ETMRs, our detailed analyses showed a range of *C19MC* alterations (Figure 1B-D) including amplification (≥10 copies) in 62.5% (90/144) and low level copy number gains (2-4 copies) in 22% (32/144) of primary ETMRs; other frequent alterations included chr 2 (64%) gains and chr 6 (25%) losses. RNAseq data available for 36 tumors showed *C19MC* fusions to the primitive ion channel locus *TTHY1* in a majority (89%) but not all ETMRs with *C19MC* copy number changes. Histopathologic review (n=149) revealed a range of overlapping histologies including ETANTR (92; 62%), Medulloepithelioma (15; 10%) and Ependymoblastoma (7; 4.6%), however, 23.4% (35) of primary ETMRs lacked classic histology and met criteria for Embryonal tumors NOS (Fig 1E). A majority of ETMRs (80%) had strong, diffuse LIN28A IHC stain, however, 20% of tumors had patchy LIN28A expression. There was no correlation of tumor histology with *C19MC* alterations or LIN28A expression pattern.

**Figure 1.**
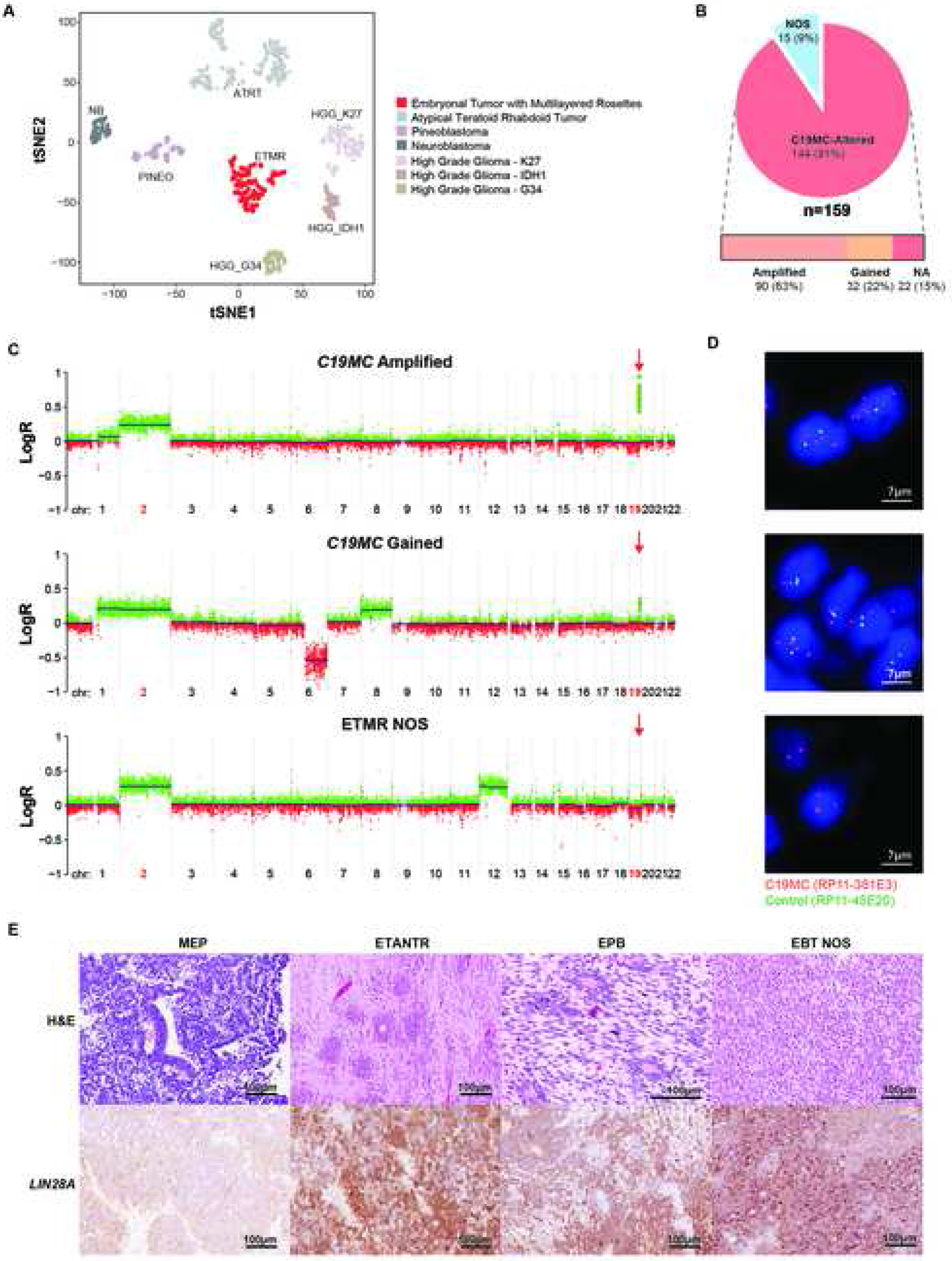
Histopathologic and molecular features of primary ETMRs. A. t-Stochastic Neighbor Embedding (tSNE) plots of DNA methylation cluster patterns of 159 ETMRs relative to a representative set (n=451) of reference pediatric CNS tumors. Tumor types are denoted as colored spheres; *C19MC*-altered ETMRs and ETMR NOS are shown in a single cluster. B. Type and frequency of *C19MC* alteration in ETMRs; NA: Not available, FISH quantification not available. C. Whole genome copy number profile of representative ETMRs with amplification (top), gain (middle), or no alteration of *C19MC* (ETMR NOS) (bottom) generated using 850K Illumina methylation array data. Copy numbers are shown relative to a logR scale for probe intensity. Chromosome locations are designated on the X-axis; arrows indicate *C19MC* map position. D. Representative cases of ETMRs analysed by fluorescence *in situ* hybridization (FISH) with *C19MC* specific (Red; RP11-381E3) and control (Green; RP11-45E20) BAC probes showing amplification (top), gain (middle), and no copy number alterations (bottom) of *C19MC*. E. Representative H and E, and LIN28A immunohistochemical (IHC) stains of ETMRs with histologic features of medulloepithelioma (MEP), Embryonal Tumors with Abundant Neuropil and True Rosettes (ETANTR), ependymoblastoma (EPB) and Embryonal Brain Tumor Not Otherwise Specified (EBT NOS).

### Clinical features of patients at diagnosis

To elucidate clinical phenotypes, we examined demographics (age, gender) and disease features (tumor location, stage) for the 159 primary ETMR patients. Unlike other pediatric brain tumors, ETMR patients exhibited a male to female ratio of 1:1.4 (Figure 2A). Median age at diagnosis was 26 months (0.5-141; IQR 18-36) with 96% of patients presenting between 12-60 months of age (Figure 2B); only 2-4% of patients were < 6 months and > 10 years of age at diagnosis. Age and gender were not significantly different for patients with *C19MC-altered* ETMRs or ETMR NOS. ETMRs arose from multiple CNS sites, most commonly in the cerebrum (55%) and cerebellum (18%); rare tumors were in midline (11%), intra-ventricular (5%), lumbo-sacral (2%) and orbital (1%) locations. Cerebral tumors arose as frontal (24%), parietal (18%), temporal (11%) and occipital (5%) lesions; 32% were large masses traversing lobes or supra-and infra-tentorial compartments. Notably, 10% of ETMRs presented as intrinsic brain stem tumors with radiologic resemblance to diffuse intrinsic pontine glioma (DIPG) seen in older children (Figure 2C-D). ETMRs were localized in 73% of patients at diagnosis (Figure 2E); rare ETMRs had extension and invasion of local structures reminiscent of mesenchymal tumors (Table S2). Metastatic diseases seen in 27% of patients at diagnosis, was frequently advanced with nodular cranio-spinal disease (62% M2-3) and rare extra-neural metastases (M4); only 15% had isolated positive (M1) CSF cytology.

**Figure 2.**
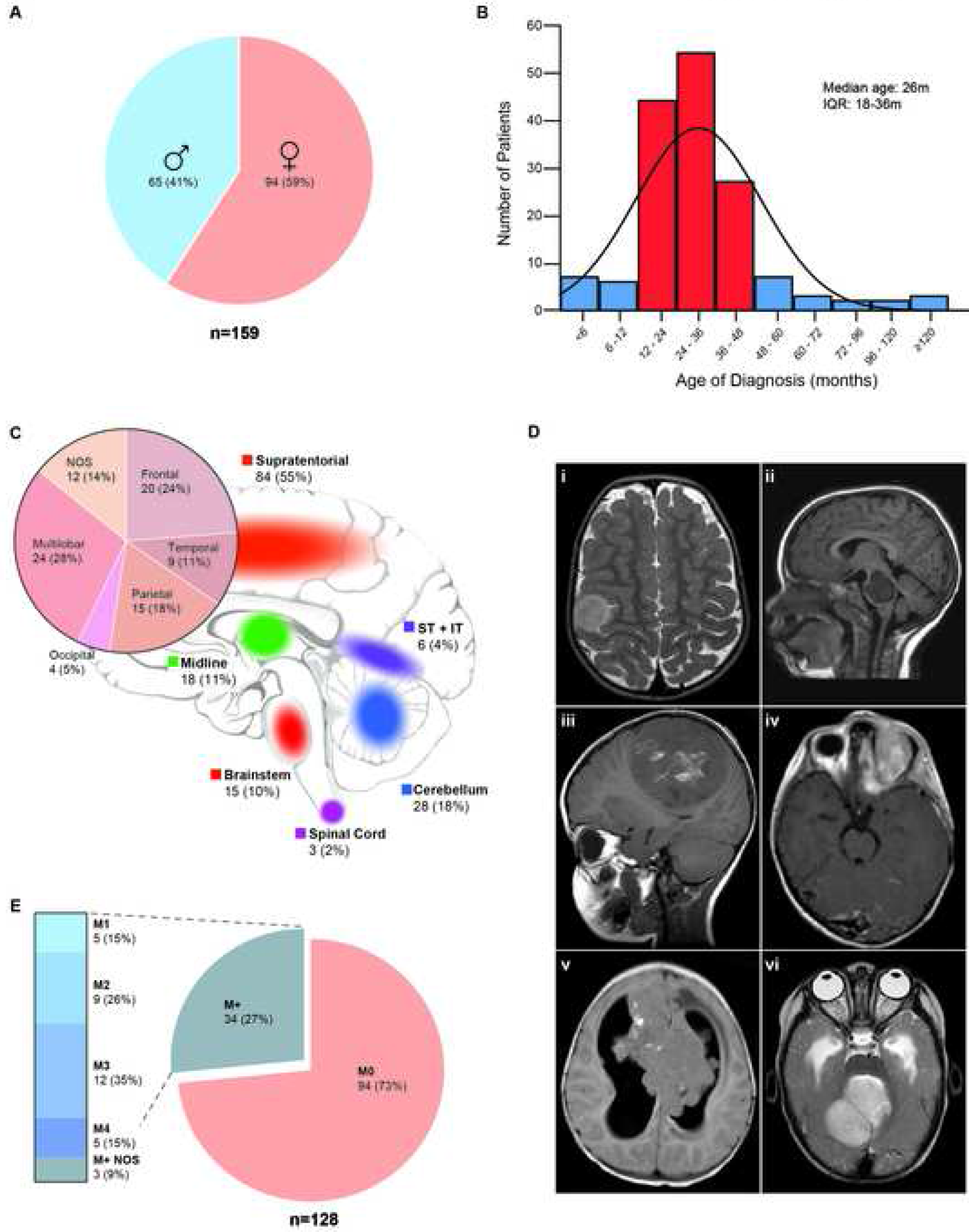
Clinical features of ETMR patients at diagnosis. A. Distribution of gender, n=159. B. Distribution of age at diagnosis for 155 patients; red bars indicate median age and range. C. Distribution of primary ETMRs in different CNS locations, n=154. D. Representative MRI images of primary ETMRs in cortical, brainstem, parietal, intra-orbital, intraventricular and cerebellar locations are shown respectively in panels i-vi. E. Distribution of disease stage as per Chang staging M0 (non-metastatic) and M+, M1-4 (metastatic) of 128 primary ETMRs.

### Disease trajectory and clinical prognostic factors

To evaluate disease patterns and prognosticators, we performed survival analysis for 108 patients with complete follow-up data after curative multi-modal regimens including surgery, chemotherapy and/or radiation. ETMR patients had respective median EFS and OS of 6 (IQR 4-15) and 13 months (IQR 7-21) (Figure 3A-B) at a median follow-up of 25 months (IQR 12-78). Survival analyses indicated ETMRs patients had rapid disease tempo with respective 6, 12 and 24-month median EFS of 57% (95% CI 47-67), 37% (95% CI 26-48) and 31% (95% CI 20-41) (Table 1). Despite multi-modal therapies, 62% (67/108) of patients succumbed within 24 months of diagnosis, with 2 and 4-year OS of 29% (95% CI 20-38) and 27% (95% CI 18-37). Univariate analyses identified brain stem location and metastatic disease at diagnosis as significant negative risk factors for OS (Table 2) with respective 2.5 (HR=0.40 95%CI 0.021-0.75, p=0.005), and 2.3 fold (HR=0.44 95% CI 0.2612-0.74, p=0.002) greater risk of death. Age, gender and *C19MC* status were not significantly associated with EFS or OS.

**Figure 3.**
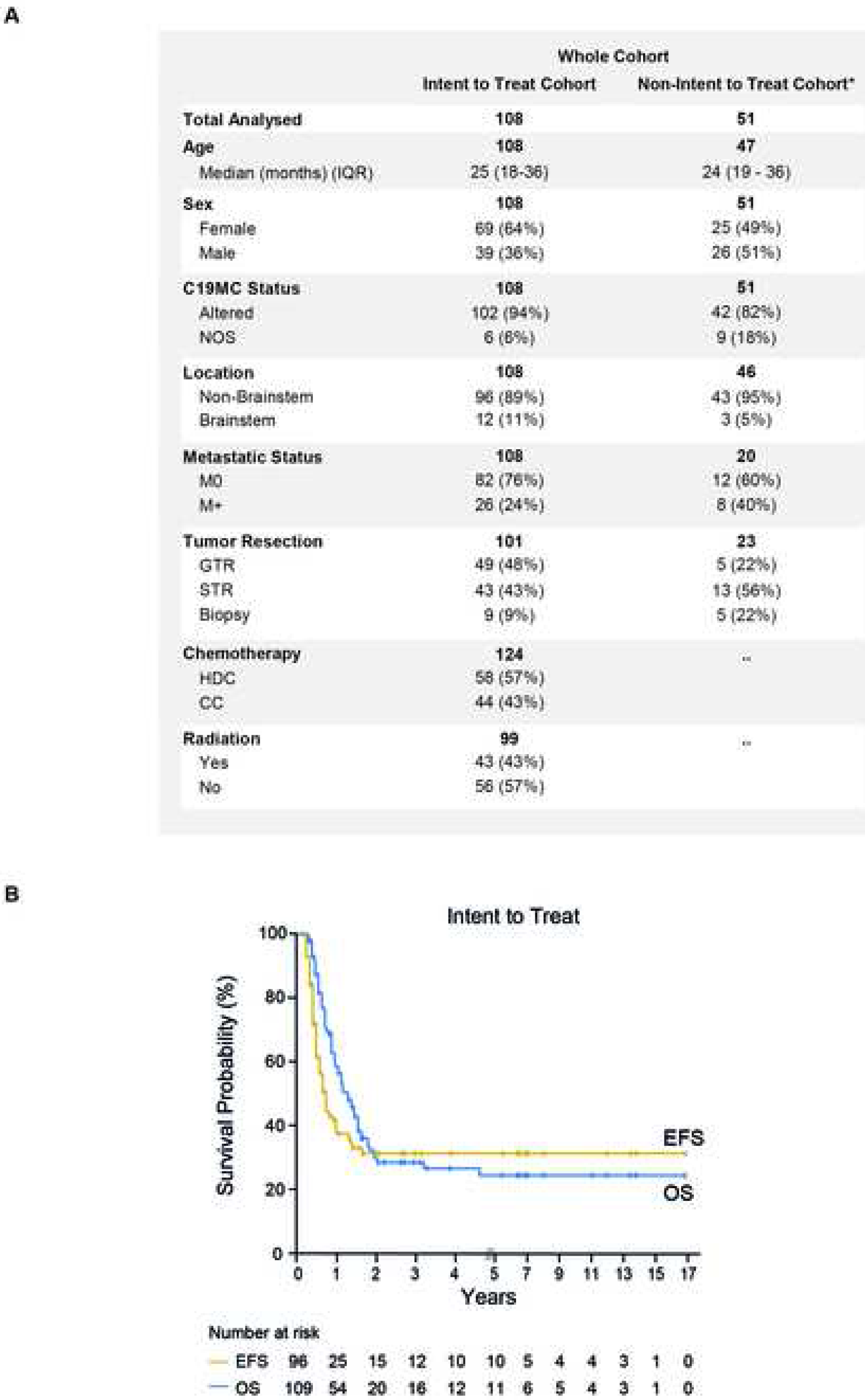
Characteristics of ETMR patients in intent to treat versus non-intent to treat cohort. A. Disease and treatment features of patients treated with (n=108) and without curative regimens (n=51). B. Event free survival (EFS) and overall survival (OS) of 108 patients treated with curative intent, using Kaplan-Meier log rank analyses.

**Table 1.** EFS and OS for patients treated with curative intent.

|  | % EFS (95% CI) |  |  |  |  |  | % OS (95% CI) |  |  |
| --- | --- | --- | --- | --- | --- | --- | --- | --- | --- |
|  | n | 3 month | 4 month | 6 month | 12 month | 24 month | n | 2 year | 4 year |
| **Entire Cohort** | 94 | 84 (77 - 91) | 72 (62 - 81) | 57 (47 - 67) | 37 (26 - 48) | 31 (20 - 41) | 107 | 29 (20-38) | 27 (18- 37) |
| **Location** | 93 |  |  |  |  |  | 107 |  |  |
| Non-Brainstem | 82 | 86 (79 - 94) | 76 (66 - 85) | 64 (53 - 74) | 42 (30 - 54) | 35 (23 - 47) | 95 | 35 (23 - 47) | 35 (23 - 47) |
| Brainstem | 11 | 69 (44 - 94) | 46 (19 - 73) | 8 (0 - 22) | 8 (0 - 22) | 8 (0 - 22) | 12 | 8 (0 - 22) | 8 (0 - 22) |
| **Metastatic Status** | 89 |  |  |  |  |  | 107 |  |  |
| M0 | 71 | 84 (76 - 92) | 70 (60 - 81) | 57 (45 - 68) | 42 (30-54) | 36 (24 - 48) | 81 | 36 (24-47) | 33 (22 - 45) |
| M+ | 18 | 82 (64 - 100) | 77 (56 - 97) | 45 (20 - 69) | 19 (4 - 34) | 7 (0 - 21) | 26 | 4 (0-13) | 4 (0 - 13) |
| **Tumor Resection** | 92 |  |  |  |  |  | 92 |  |  |
| GTR | 49 | 93 (86 - 100) | 87 (77 - 96) | 72 (60 - 85) | 54 (39 - 70) | 48 (32 - 64) | 49 | 44 (29-59) | 40 (24 - 56) |
| STR | 43 | 77 (64 - 90) | 63 (48 - 79) | 40 (24 - 56) | 24 (9 - 39) | 17 (4 - 30) | 43 | 21 (8-21) | 14 (0 - 28) |
| **Chemotherapy** | 92 |  |  |  |  |  | 102 |  |  |
| CC | 36 | 64 (48 - 80) | 51 (34 - 68) | 38 (21 - 54) | 24 (9 - 39) | 15 (2 - 29) | 44 | 18 (5-30) | 11 (0 - 21) |
| HDC | 56 | 96 (91 - 100) | 85 (75 - 95) | 70 (57 - 82) | 46 (32 - 60) | 44 (27 - 55) | 58 | 38 (24-52) | 38 (24 - 52) |
| **Radiation** | 91 |  |  |  |  |  | 99 |  |  |
| Yes | 40 | 97 (90 - 104) | 84 (72 - 96) | 76 (63 - 90) | 56 (39 - 72) | 49 (32 - 66) | 43 | 43 (27-59) | 43 (27 - 59) |
| No | 51 | 88 (79 - 97) | 88 (79 - 97) | 38 (24 - 52) | 21 (9 - 33) | 14 (3 - 26) | 56 | 21 (9-33) | 14 (3 - 25) |
| **C19MC Status** | 89 |  |  |  |  |  | 107 |  |  |
| Altered | 84 | 83 (76 - 91) | 70 (61 - 80) | 56 (45 -66) | 35 (24 - 46) | 30 (20 - 41) | 101 | 26 (17 - 36) | 26 (17 - 36) |
| NOS | 5 | 100 | 100 | 75 (32 - 100) | 66 (32 - 100) | 50 (1 - 99) | 6 | 33 (0 - 83) | 33 (0 - 83) |

EFS: Event Free Survival, OS: Overall Survival, GTR: Gross Total Resection, STR: Subtotal Resection, HDC: High Dose Chemotherapy, CC: Conventional Chemotherapy

### Treatment related prognostic factors

As best treatment approach for ETMRs remains unknown, we investigated the impact of conventional embryonal brain tumor modalities in our cohort (Table 2). Amongst the 108 patients treated with curative intent, 48% had complete tumor resection (GTR), 57% received high dose chemotherapy (HDC) and 43% received radiation (56% focal; 40% cranio-spinal) as part of primary tumor treatment. Univariate hazard analyses identified receipt of GTR (HR=0.43; 95% CI 0.25-0.72, p= 0.002), HDC (HR= 0.41 95% CI 0.25-0.67, p <0.001) and radiation (HR=0.48 95% CI 0.25-0.81, p = 0.005) as significant positive prognosticators for OS. In multi-variable analyses all three modalities also correlated significantly with EFS and OS. GTR was associated respectively with 55% and 62% risk reduction in EFS (HR=0.45 95% CI 0.25-0.81, p= 0.01) and OS (HR=0.38 95% CI 0.21-0.68, p= 0.001), while HDC treatment also correlated with superior EFS (HR=0.45 95% CI 0.25-0.81, p=0.01) and OS (HR=0.55 95% CI 0.31-0.97, p= 0.04). Notably, focal or cranio-spinal radiation was associated with a 77% (HR=0.23 95% CI 0.12-0.47, p <0.001) and 68% (HR=0.32 95% CI 0.16-0.60, p<0.001) respective risk reduction in EFS and OS. These data indicate ETMR patients benefit from a combination of all three conventional treatment modalities.

**Table 2.** COX proportional Hazard Analyses of clinical and treatment variables.

| Variable | Subjects | Hazard Ratio (95% CI) | p value |
| --- | --- | --- | --- |
| **Univariate OS** |  |  |  |
| Sex (male vs female) | 107 | 0.82 (0.50-1.33) | 0.43 |
| Age (continuous) | 107 | 0.99 (0.98-1.00) | 0.42 |
| All Locations | 108 | 0.83 (0.74-0.93) | 0.002 |
| Non-Brainstem vs Brainstem | 107 | 0.40 (0.21-0.75) | 0.005 |
| Metastatic Status (M0 vs M+) | 107 | 0.44 (0.26-0.74) | 0.002 |
| C19MC (Altered vs NOS) | 107 | 0.40 (0.09-1.66) | 0.21 |
| Tumor Resection (GTR vs STR) | 92 | 0.43 (0.25-0.72) | 0.002 |
| Chemotherapy (HDC vs CC) | 102 | 0.41 (0.25-0.67) | <0.001 |
| Radiation (yes vs no) | 99 | 0.48 (0.25-0.81) | 0.005 |
| **Multiple Variable EFS** |  |  |  |
|  | **78** |  |  |
| Tumour Resection (GTR vs STR) | 43 : 35 | 0.45 (0.25-0.81) | 0.01 |
| Chemotherapy (HDC vs CC) | 49 : 29 | 0.45 (0.25-0.81) | 0.01 |
| Radiation (yes vs no) | 32 : 46 | 0.23 (0.12-0.47) | <0.001 |
| **Multiple Variable OS** |  |  |  |
|  | **80** |  |  |
| Tumour Resection (GTR vs STR) | 44 : 36 | 0.38 (0.21-0.68) | 0.001 |
| Chemotherapy (HDC vs CC) | 50 : 30 | 0.55 (0.31-0.97) | 0.04 |
| Radiation (yes vs no) | 34 : 46 | 0.32 (0.16-0.60) | <0.001 |

EFS: Event Free Survival, OS: Overall Survival, GTR: Gross Total Resection, STR: Subtotal Resection, HDC: High Dose Chemotherapy, CC: Conventional Chemotherapy

### Multi-modality therapy confers survival advantage in ETMR patients

Due to severe neuro-cognitive toxicity of whole brain and spine radiation in younger children, post-surgical HDC regimen is often used as a radiation deferral or avoidance strategy for younger children with embryonal brain tumors. We investigated the relative importance of surgery, intensity of chemotherapy and radiation to primary disease control and survival in ETMR patients. All patients irrespective of disease stage and location were stratified by extent of surgery, HDC versus CC, and radiation-free or primary radiation treatment (Figure 4 A-D). Log-rank analyses showed HDC treatment significantly but nominally improved EFS and OS (10%±9%; p=0.001 and 0.005) for patients with STR, while patients with GTR had superior but non-significant improvement in EFS (37±14%) and OS (32±13%) as compared to EFS and OS of 0% for all patients treated with only CC (Figure 4A-B). These findings suggest HDC intensification improves disease control and survival for ETMR patients, and that a proportion of patients with GTR may be cured with HDC alone.

**Figure 4.**
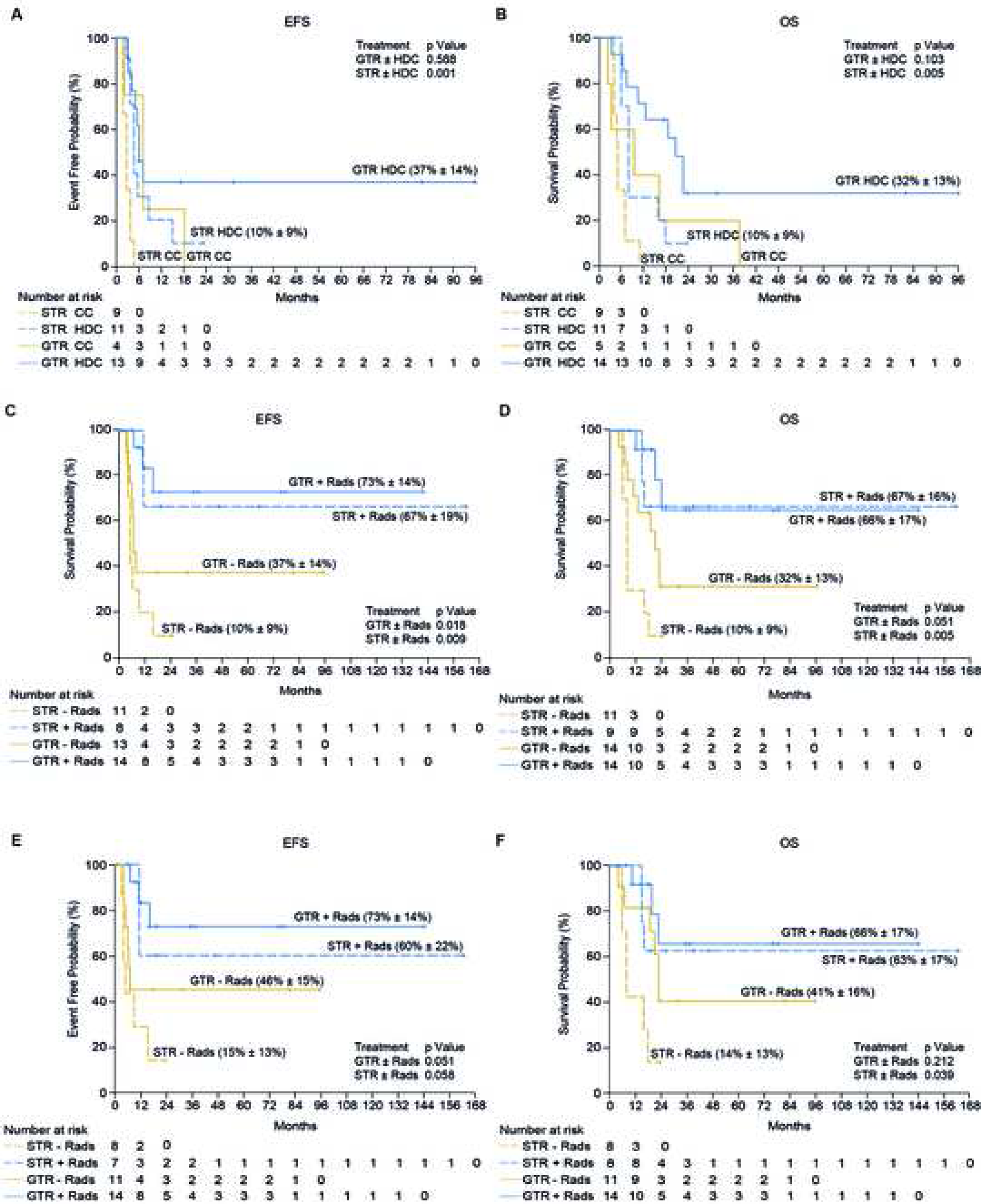
Impact of treatment modalities on ETMR patient survival. Log rank analyses of Event Free (EFS) and Overall survival (OS) for 108 patients with primary ETMRs treated with curative intent stratified by extent of tumor resection (GTR versus STR), receipt of high dose chemotherapy (HDC) or conventional chemotherapy (CC) and radiation (+ Rads) or no radiation (- Rads). A-B. EFS (n= 37) and OS (n= 39) patients treated with only HDC and CC and no radiation. C-D. EFS (n= 46) and OS (n= 48) for all patients treated with HDC, stratified by extent of surgery and radiation. E-F. EFS (n= 40) and OS (n= 41) for patients with localized, non-brain stem ETMR treated with HDC, stratified by extent of surgery and radiation.

To examine if radiotherapy conferred additional benefit in patients treated with HDC, we performed log rank analyses for patients treated with HDC stratified by receipt of radiation and extent of surgery. Consistent with multi-variable analyses, patients with STR treated with HDC and radiation had significantly improved EFS and OS, respectively 67±19% and 67±16% as compared to 10±9% for both EFS and OS (p=0.009; p=0.05) in non-radiated counterparts (Figure 4C-D). Disease control in patients with GTR treated with HDC and radiation was also significantly superior with EFS of 73 ±14% compared to 37±14% for non-radiated patients (p=0.018). However, we observed a strong but non-significant trend in OS, respectively 66±17% and 32±13% for patients with GTR treated with and without radiation (p=0.051). We next examined effects of HDC and radiation in patients with lower risk clinical features as identified in uni-variate analyses (M0, non-brain stem primaries). These analyses showed patients with STR treated with HDC and radiation also had better OS of 63 ± 17% versus 14±13% in non-radiated counterparts (p=0.039). Interestingly, similar to the global cohort, radiation correlated with a strong but non-significant trend towards improved EFS (73 ±14% vs 46%±15%; p=0.051) and OS (66±17% and 41 ± 16%; p=0.212) for patients with GTR (Figure 4E-F).

Consistent with the importance of intensified multimodal regimens in ETMR survival, we observed 52% of patients with OS greater than the cohort median of 13 months received primary RT as compared to 25% of patients with EFS less than the cohort median of 6 months, (range 1-6 months; p=0.01). Similarly, 70% and 30% of patients with OS ≥ 13 months (range 13-165 months) respectively received HDC and CC treatment (p=0.04) (Table 3). While our aggregate data indicate surgery, HDC and radiation have important and inter-dependent roles in ETMR survival, it is notable that 24% of 21 long-term survivors (OS >24 −102 months) received only HDC and no radiation, suggesting use and timing of radiation may be further tailored to patient-specific risk features.

**Table 3.** Clinical and treatment characteristics of patients with EFS < 6 months and OS > 13 months.

|  | EFS ≤ 6 months | OS ≥ 13 months | p value |
| --- | --- | --- | --- |
| Number of patients | 48 | 56 |  |
| Median Age (months) | 24 | 25 |  |
| Gender |  |  |  |
| Female | 29 (60%) | 36 (64%) | 0.69 |
| Male | 19 (40%) | 20 (36%) |  |
| Tumour Location |  |  |  |
| Non-Brainstem | 38 (79%) | 52 (93%) | 0.05 |
| Brainstem | 10 (21%) | 4 (7%) |  |
| Metastasis |  |  |  |
| Yes | 11 (23%) | 9 (16%) | 0.31 |
| No | 37 (77%) | 47 (84%) |  |
| Tumor Resection |  |  |  |
| GTR | 13 (32%) | 35 (66%) | 0.05 |
| STR | 28 (68%) | 18 (34%) |  |
| Chemotherapy |  |  |  |
| HDC | 22 (49%) | 37 (70%) | 0.04 |
| CC | 23 (51%) | 16 (30%) |  |
| Radiation |  |  |  |
| Yes | 12 (35%) | 29 (63%) | 0.01 |
| No | 22 (65%) | 17 (37%) |  |
| Survival Status |  |  |  |
| Alive | 11 (23%) | 29 (52%) | 0.01 |
| Dead | 37 (77%) | 27 (48%) |  |
| Median EFS (months) | 4 (1-6) | 15 (1-165) |  |
| Median OS (months) | 7 (3-132) | 22 (13-165) |  |

EFS: Event Free Survival, OS: Overall Survival, GTR: Gross Total Resection, STR: Subtotal Resection, HDC: High Dose Chemotherapy, CC: Conventional Chemotherapy

## DISCUSSION

Through a first integrated clinical and molecular study of a substantial global registry based cohort, we describe a detailed clinical landscape for ETMRs - a rare tumor only recently designated as a new WHO diagnostic entity (ICD-O code (9478/3*) (2). We demonstrate these highly aggressive tumors in younger children, have pleomorphic histologic features and a wider clinical spectrum than previously appreciated. Importantly despite historical data indicating futility of therapy, our study shows these rapidly progressive tumors are curable with timely intensified multimodal therapy, underscoring critical, prompt accurate diagnosis for this rare disease. Our study identifies disease stage, tumor location, extent of surgery, receipt of HDC and primary radiotherapy as important prognosticators of survival. In addition to providing a much-needed framework for future clinical trials, our comprehensive and unique study of a large cohort of primary ETMRs has immediate practical implications for diagnostic and therapeutic approaches to children with this orphan disease. Although ~200 cases of ETMRs are reported to date (12, 14-18), the incidence of this disease is likely underestimated due to lack of molecular confirmation in most published studies, and reporting of ETMRs as historical histologic diagnostic categories. Furthermore, our analyses based on a large cohort with matched molecular and clinical data indicate a much wider clinical spectrum of ETMRs than previously recognized. Indeed, while ETMR is classically considered as large “supra-tentorial” tumor with frequent metastases, our data indicate 45% of primary ETMRs arise in sites characteristic of other malignant brain tumors. Furthermore rare primary ETMRs mimic benign ophthalmic lesions (19) as well as features of non-CNS malignant tumors with local extension/invasion and extra-neural metastases (Table S2). Our detailed diagnostic verification also confirm significant variation in histo-morphologic features of primary ETMRs with 15-20% lacking classic rosette-forming histology. Furthermore, amongst the 91% with *C19MC* alterations, nearly 40% of tumors had either small areas *ofC19MC* amplification, or low level *C19MC* gains evident only by FISH analyses while 10% lacked *C19MC* gene fusions by RNA sequencing assays. Together with prior clinical reports (11, 20, 21), our data indicates delayed clinical recognition and non-uniform diagnostic methods may contribute to the poor outcome of ETMRs which have distinctly rapid disease tempo. As *C19MC* genomic alterations is seen only in ETMRs and no other pediatric brain tumors (6), we propose that *C19MC* FISH together with histopathologic analyses and LIN28A immuno-stains, tools available in most pathology laboratories worldwide, be routinely performed for young patients with embryonal brain tumors to enable prompt clinical recognition and intervention.

Cumulative studies of ETMR patients diagnosed with various methods, have shown dismal outcomes with two larger series reporting short PFS and OS (15% 3yr OS 10.7 month) (8, 15). Our analyses over a more contemporary era show an overall modest improvement in ETMR survival (37% 1 year EFS; 27% 4 year OS). Importantly, we also observed remarkable EFS of 73% and OS of 66% in patients with localized, non-brain stem tumors, indicating a majority of ETMR patients with these good clinical risk features can be cured after gross total tumor removal and treatment with high dose chemotherapy and radiation.

Consistent with prior observations (15, 16, 22), radiation emerged as a strong treatment-related prognostic factor in our study. Notably, we observed that 62% of long-term survivors in our study received focal (38%) or no (24%) radiation treatment (Table S4) indicating most ETMR patients can be spared cranio-spinal radiation, and further that radiation application and timing may be tailored to patient specific risk features including extent of surgery and response to adjuvant chemotherapy. Our retrospective study lacks data on radiation timing, however, it is notable that patients with GTR treated with HDC and radiation had superior EFS but comparable 4yr OS relative to patients with STR (Figure 4C-D). Furthermore, limited data on relapses in patient with GTR post-HDC treatment alone indicate a majority (8/10) recurred locally, some multiple times. These observations suggest differences in radiation timing may contribute to disease progression and underlie observed discrepant EFS and OS in patients with GTR treated with HDC and radiation seen in our study (Figure 4E-F).

Similar to smaller reports (14-16), HDC emerged as a positive prognostic factor in our study while treatment with CC correlated with rapid progression in all patients. We observed an OS of 41 ±16%, amongst patients with M0, GTR, non-brain stem tumors treated with HDC alone while EFS and OS of radiated counterparts were not significantly different and suggest HDC may spare radiation in a significant proportion of patients with these favorable clinical features. Whether HDC has a cytoreductive role for sub-totally resected ETMRs, as reported for other embryonal tumors (23), is less clear as patients with STR but other favorable risk features had only nominally improved survival with HDC treatment alone. Our data lacked power to directly compare effects of HDC or CC combined with radiation, however, it is notable that no patients with STR treated with CC and radiation survived (0/5). Whether efficacy of CC or HDC combined with focal radiation are comparable in patients with favorable risk features could not be assessed in our study as details of radiation were only available for 2/5 long-term survivors treated with CC.

Fifty two percent (48/92) of patients in our study had rapid (EFS 1-6 months) local and/or disseminated progression on therapy. Patients with brain stem and/or metastatic disease represented the highest risk groups; 9/10 patients with biopsied or partially resected brainstem primaries progressed at a median of 4 months despite HDC and radiation. A sole survivor (OS=202 months) had GTR, indicating well demarcated brainstem tumors may benefit from maximal safe surgery (24). Of remark, 27% of patients with early local recurrence had favorable risk features, and suggest invasive tumor biology contributes to early local tumor regrowth in a proportion of ETMRs patients. Of note, a number of these are long-term survivors after second surgery and/or focal radiation (Table S4) indicating prompt focal radiation may particularly benefit patients with invasive tumors, or early local recurrence.

To date ETMRs have been considered highly fatal diseases; no curative therapy was undertaken for 9% of patients, including those with localized disease, in our registry Notwithstanding the inherent limitations of retrospective studies, our collective data indicates ETMR is an imminently curable disease and suggest prompt diagnosis and intervention with risk tailored multi-modal therapy can cure a significant proportion of patients. We propose that maximum safe surgery followed by HDC be undertaken for children presenting with localized ETMRs, and second-look surgery and/or early radiation be considered for children with recurrence post complete surgery or while on chemotherapy treatment. Experimental approaches or agents are needed for children with advanced metastatic or progressive disease, but these must be considered within the context of a clinical trial with accompanying bio-marker studies to further inform biological and risk stratified therapies. While effective, treatment of the very young ETMR patients with multi-agent chemotherapy, radical surgery and even limited field radiation has long-term physiologic and neuro-cognitive implications. Thus, it will be important to delineate the molecular underpinnings of invasive biology and treatment failure in ETMRs to develop new biological therapies with less toxicity. However innovative therapeutics for ETMR remain limited due to lack of animal models and human tumor cell lines available for study. How *DICER 1* germline alterations, reported in a small number of patients, contribute to ETMR phenotypes or therapeutic response remains to be studied (25-27). Nonetheless, recent studies suggest epigenetic mechanisms and defective DNA damage repair represent important vulnerabilities in ETMRs (28). Studies to date identify inhibitors of mTOR, Topoisomerases, PARP, Aurora and Polo-kinases, Histone Deacetylases, Bromodomain proteins and DNA methylation (Vorinostat, 5AzaC) as promising novel agents (26, 28-30).

Despite availability of a specific molecular marker for ETMRs, there remains significant gaps in diagnostic and treatment approaches to this aggressive disease. Indeed, our registry data indicate only a small proportion of ETMR patients have molecularly established diagnosis while CSF analyses, critical for robust staging of embryonal brain tumors, are not uniformly performed. Our integrated characterization of a unique, large global registry based cohort provides a first comprehensive description of the clinical and diagnostic features of this new disease and provides an important framework for risk-tailored, practical clinical management of ETMR patients for which there is presently no established standard of care or clinical trials.

In addition to immediate clinical implications of the current study, the extensive global clinical registry and biorepository of the RBTC network will continue to be a critical resource for future clinical and biological studies. Thus, it is vital we continue to enroll children diagnosed with ETMR and other rare tumors in the Global Rare Brain Tumor Consortium Clinical Registry and Biorepository (Rarebraintumorconsortium.ca).

### Research in Context

ETMR is a rare, highly malignant brain tumor, previously considered as various separate histologic diagnoses, and only recently designated as a distinct WHO diagnostic entity based on cumulative molecular studies. Early reports described ETMRs as frequently disseminated and futile diseases, which has led to increasing use of upfront experimental therapies in these patients. However as well annotated molecular and clinical data on a large cohort of ETMR is lacking, the full spectrum of ETMRs clinical phenotypes and the impact of conventional treatment modalities on ETMR patient survival has remained poorly understood. Our comprehensive study of a large global registry based cohort show ETMRs are primarily with localized diseases and that aggressive intervention with maximum safe surgery, high dose chemotherapy combined with timely radiation provides significant long-term survival benefits for ETMR patients. Our analyses show ETMRs share overlapping clinical features with other childhood cancers, but have distinctly rapid disease tempo thus routine use of specific molecular assays for ETMRs is critical to avoid delays in diagnosis and treatment. We identify key clinical prognosticators and therapeutic modalities that are important for construction of future risk-stratified prospective trials. In addition, our study provides a critical framework for clinical and diagnostic work-up, as well therapeutic approaches to ETMRs and thus have immediate clinical practice implications for children diagnosed worldwide with this rare disease.

## Data Availability

Supplemental data will be publicly available pending publication.

## FIGURE LEGENDS

**Consort Figure.**
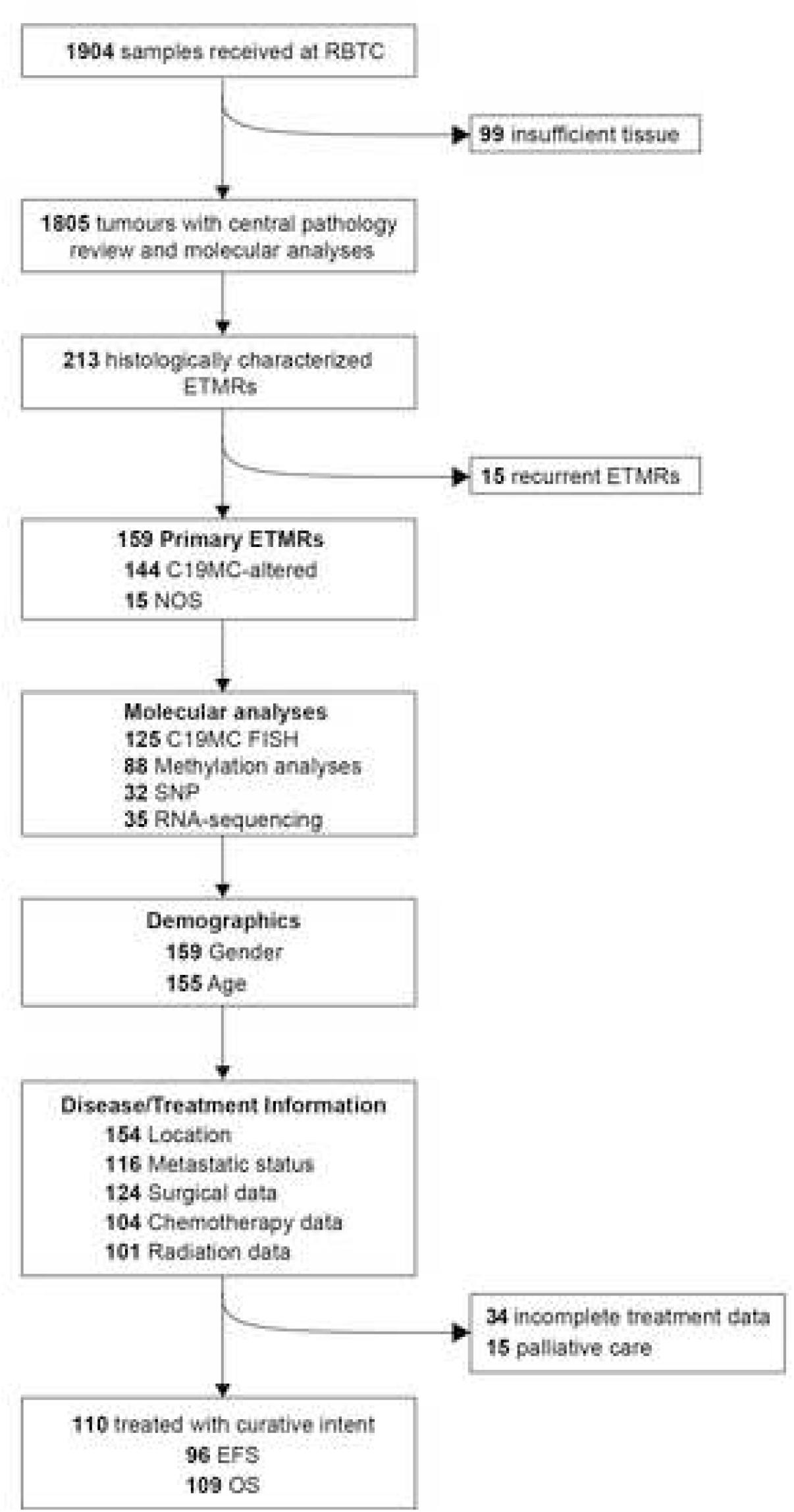
Identification and analyses of primary ETMRs from the Rare Brain Tumor Consortium Clinical Registry and Biorepository.

